# Incidence of SARS-CoV-2 infection in a cohort of workers from the University of Porto

**DOI:** 10.1101/2021.10.14.21264980

**Authors:** Joana Pinto Costa, Paula Meireles, Pedro N. S. Rodrigues, Henrique Barros

**Author notes:** **Correspondence to:** Joana Pinto da Costa, Address: Instituto de Saúde Pública da Universidade do Porto, Rua das Taipas, n° 135 4050-600, Porto, Portugal.

## Abstract

**Background:** Repeated serosurveys in the same population provide more accurate estimates of the frequency of SARS-CoV-2 infection and more comparable data than notified cases. We aimed to estimate the incidence of SARS-CoV-2 infection, identify associated risk factors, and assess time trends in the ratio of serological/molecular diagnosis in a cohort of university workers.

**Methods:** Participants had a serological rapid test for SARS-CoV-2 Immunoglobulins M and G, and completed a questionnaire, in May-July 2020 (n=3628) and November 2020–January 2021 (n=2661); 1960 participated in both evaluations and provided data to compute the incidence proportion and the incident rate. Crude and adjusted incidence rate ratios (aIRR) and 95% confidence intervals (CI) were computed using generalised linear models with Poisson regression.

**Results:** The incidence rate was 1.8/100 person-month (95%CI 1.6-2.1), and the 6 months’ cumulative incidence was 10.7%. The serological/molecular diagnosis ratio was 10:1 in the first evaluation and 3:1 in the second. Considering newly identified seropositive cases at the first (n=69) and second evaluation (n=202), 29.0% and 9.4% never reported symptoms, respectively, 14.5% and 33.3% reported contact with a confirmed case and 82.6%, and 46.0% never had a molecular test. Males (aIRR: 0.59; 95%CI: 0.42-0.83) and “high-skilled white-collar” workers (aIRR: 0.73, 95%CI: 0.52-1.02) had lower incidence of infection.

**Conclusion:** University workers presented a high SARS-CoV-2 incidence while restrictive measures were in place. The time decrease in the proportion of undiagnosed cases reflected the increased access to testing, but opportunities continued to be missed, even in the presence of COVID-19 like symptoms.

**What is already known on this subject:**

- The median ratio of seroprevalence to the corresponding cumulative incidence is 18, however, there is great variability between studies.
- Seroprevalence studies are essential to estimate the true burden of the infection.
- Few cohort studies focused on essential non-healthcare workers, such as university workers.

**What this study adds:**

- This longitudinal seroprevalence study among university workers found a SARS-CoV-2-specific IgM or IgG incidence rate of 1.8/100 person-month, and a 6 months’ cumulative incidence of 10.7%.
- The undiagnosed fraction was 3:1 in the second evaluation, representing a decrease from a 10:1 in the first evaluation in the same population showing that a gap to test-trace-isolate remained in this highly educated working population.
- Seropositive participants were mostly pauci- or symptomatic with no known contact with a COVID-19 confirmed case; “high-skilled white-collar” workers were at lower risk of being an incident seropositive case.

## Introduction

During the coronavirus disease 2019 (COVID-19) pandemic, repeated serological tests in the same population are recommended by the World Health Organization (WHO) (1). It allows a better characterization of the evolution of the severe acute respiratory syndrome coronavirus 2 (SARS-CoV-2) infection and the dynamics of seroconversion, either due to past infection or vaccination. Repeated serosurveys in the same population provide a more accurate estimate of the extent of SARS-CoV-2 infection and more comparable data over time than just notified cases, which are dependent on case definition, testing capacity, and testing criteria in place (2). Several seroprevalence studies have demonstrated that the extent of the infection is much larger than based on molecular diagnosis (3–9). A study estimated the median ratio of seroprevalence to cumulative incidence to be approximately 18 (9). However, repeated serosurveys in the same population cohort are uncommon, essentially encompassing recovered patients and healthcare workers (10–12). Moreover, few studies addressed higher education workers - mainly assessed using cross-sectional serosurveys early in the pandemic (13–16).

In Portugal, the first case of COVID-19 was diagnosed on March 2, 2020. The first national serological survey (ISN COVID-19), conducted between May and July 2020 in a sample of 2301 inhabitants provided a seroprevalence of 2.9% (4) and a second one, conducted from February to March 2021, estimated a 13.5% prevalence, excluding those already vaccinated (17).

While there were no preventive pharmaceutical measures available, mitigation strategies were based on physical distance, facial masks, or handwashing measures and have led to the closure of schools around the world in an unprecedented disruption to global education systems. In Portugal, schools at all levels were closed on March 16, 2020, and remote working was mandatory from March 18 to June 30, 2020. Schools reopened at the start of the 2020/2021 academic year, mid-September 2020, up to mid-January 2021. An initial seroprevalence study of 4592 workers of all public higher education institutions of Porto was conducted between May and July 2020 and captured the seroprevalence when lockdown measures were eased (15). A follow-up seroprevalence study – from November 2020 to January 2021 - among workers from the University of Porto (U.Porto) was conducted to capture the evolution of the infection in this community. In this study, we aimed to estimate the incidence of SARS-CoV-2 infection in a non-vaccinated group, to identify factors associated with SARS-CoV-2 incidence, and the time trends in the ratio between serological evidence and a reported molecular diagnosis.

## Methods

All the workers of the U.Porto were invited to participate in two serological surveys using a point of care test for SARS-CoV-2 specific IgM and IgG antibodies. The first evaluation occurred from May 21 to July 31, 2020, and the second from November 27, 2020, to January 29, 2021. Participation was voluntary, and scheduling was initiated by the workers. The first evaluation is described in detail elsewhere (15).

In both evaluations data were collected by trained researchers, using a structured questionnaire including information on sex, age, nationality, comorbidities (defined as having a disease that requires regular medical care e.g. treatments, appointments, etc.), the highest level of education completed, profession (categorized in occupation classification and dichotomized in “high-skilled white-collar”: yes or no), remote working at the time of testing, county of residence [classified according to the Nomenclature of Territorial Units for Statistics Level 3 (NUTS 3) and aggregated in Metropolitan area of Porto (MAP) and Outside MAP], and self-perceived probability of having been infected. Also included infection related questions: contact with a confirmed SARS-CoV-2 case and quarantine, symptoms since the beginning of 2020 (categorized into asymptomatic; paucisymptomatic: one or two of the following symptoms: cough, dyspnea, odynophagia, headache, vomiting, or nausea, diarrhea, asthenia, or fever; and symptomatic: at least three of the listed symptoms, or dysgeusia or anosmia) and previous SARS-CoV-2 diagnostic tests and diagnosis.

Participants provided written informed consent to all procedures. The study protocol was approved by the ethics committee of the Institute of Public Health of the University of Porto (ID 20154) and all procedures complied with the principles embodied in the Declaration of Helsinki.

### SARS-CoV-2 specific IgM and IgG antibodies determination and follow-up

During the first evaluation two point-of-care tests were used – the STANDARD Q COVID-19 IgM/IgG Duo used from May 21 to July 9, n=3040 (manufacturer reported sensitivity of 92.6% eight days after symptom onset and specificity of 96.5% for both IgG and IgM); and the STANDARD Q COVID-19 IgM/IgG Combo from July 10 to July 31, n=588 (manufacturer reported sensitivity of 94.5% seven or more days after symptom onset and specificity of 95.7% for both IgG and IgM). In the second evaluation, only the STANDARD Q COVID-19 IgM/IgG Combo was used (n=2661).

### Statistical analysis

An incident case was defined as having a reactive result either for IgM or IgG in the second evaluation, among those who participated in both evaluations and were seronegative in the first. To calculate the incident rate, time at risk was computed as the time in months between the two serological tests in case of seronegative individuals, for the incident cases without a previous diagnosis we considered half time between the two tests, and for those with a molecular diagnosis, we considered the time between the first serological test and the date of molecular diagnosis. Associations were described using crude and adjusted incidence rate ratios (aIRR) and respective 95% confidence intervals (CI), adjustments were made for sex, age (continuous), nationality, and occupation classification computed using generalised linear models with Poisson regression, with the default log link and offset in the variable time at risk. A bilateral significance level of 5% was considered. Analysis was performed using IBM SPSS Statistics for Windows, Version 27.0. Armonk, NY: IBM Corp.

## Results

In total, 4329 individuals participated in the serosurveys: 3628 in the first evaluation and 2661 in the second; 1960 of those participated in both. The seroprevalence of SARS-CoV-2 infection increased from 4.1% (150/3628) in the first evaluation to 13.2% (350/2661) in the second; based on a reported previous molecular diagnosis the prevalence of the infection increased from 0.4% (16/3628) to 4.4% (118/2661), respectively. The ratio of workers who were seropositive to those with a previous molecular diagnosis was 10 to 1 in the first evaluation and 3 to 1 in the second.

Considering the 1960 workers who participated in both evaluations, 271 (13.8%) were ever seropositive, 69 (3.5%) were seropositive in the first and 251 (12.8%) were seropositive in the second evaluation (Table S1). Of the 1891 workers that participated in both evaluations and were seronegative at the first, 202 (10.7%) became seropositive – incident cases – over 11,222 person-months of observation. The overall incidence rate (IR) was 1.8 per 100 person-months (95%CI: 1.6-2.1). Additionally, among the 69 seropositive (IgM and/or IgG) cases identified in the first evaluation 20 (29.0%) were seronegative in the second, and of the 18 IgG-only reactive at the first evaluation, 7 (38.9%) were IgM and IgG reactive at the second; the detailed results according to the class of antibodies are presented in Table S1. Of the 1960 workers, 77 (3.9%) ever reported a positive molecular diagnosis. Of the 1940 workers without a positive RT-PCR at first evaluation, 70 (3.6%) had a molecular diagnosis during the follow-up period.

The distribution of incident cases is presented in Table 1. Males (aIRR: 0.59, 95%CI: 0.42-0.83) and workers classified as “high-skilled white-collar” (aIRR: 0.73, 95%CI: 0.52-1.02) were at lower risk of infection. Also, it is worth mentioning that those with basic education and living in the Porto metropolitan area had a 42% and 58% increase in incidence, respectively, thought CI included the unit. Age strata, nationality, comorbidities, remote working at the time of the evaluation, or the self-perceived probability of infection were not significantly associated with the risk of infection.

**Table 1.**
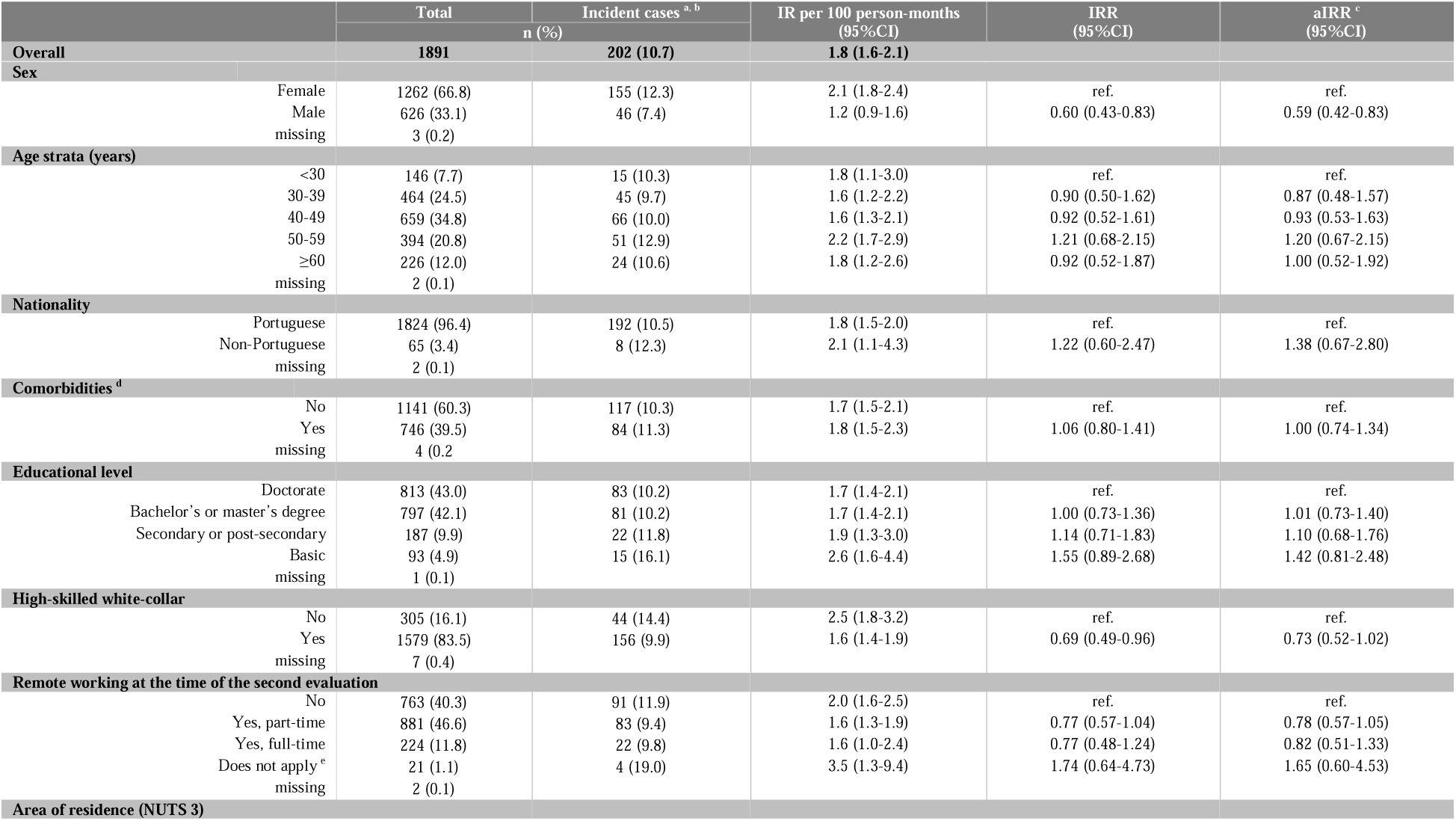
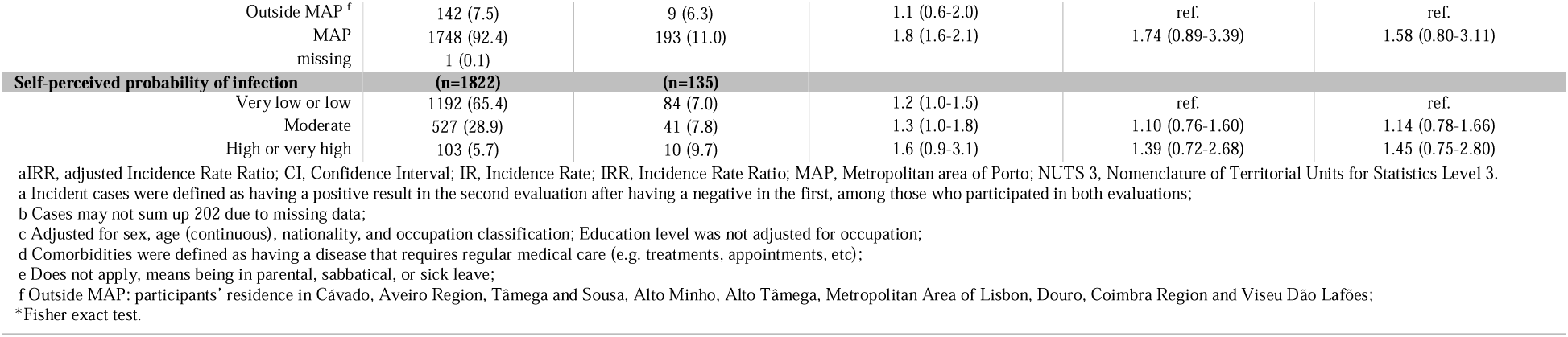
Description of the 1891 participants who took part in both evaluations and were seronegative at the first, distribution of incident cases, and factors associated with the incidence of SARS-CoV-2.

|  |  | Total | Incident cases <sup>a, b</sup> | IR per 100 person-months<br>(95%CI) | IRR<br>(95%CI) | aIRR <sup>c</sup><br>(95%CI) |
| --- | --- | --- | --- | --- | --- | --- |
|  |  | n (%) |  |  |  |  |
| **Overall** |  | **1891** | **202 (10.7)** | **1.8 (1.6-2.1)** |  |  |
| **Sex** |  |  |  |  |  |  |
|  | Female | 1262 (66.8) | 155 (12.3) | 2.1 (1.8-2.4) | ref. | ref. |
|  | Male | 626 (33.1) | 46 (7.4) | 1.2 (0.9-1.6) | 0.60 (0.43-0.83) | 0.59 (0.42-0.83) |
|  | missing | 3 (0.2) |  |  |  |  |
| **Age strata (years)** |  |  |  |  |  |  |
|  | <30 | 146 (7.7) | 15 (10.3) | 1.8 (1.1-3.0) | ref. | ref. |
|  | 30-39 | 464 (24.5) | 45 (9.7) | 1.6 (1.2-2.2) | 0.90 (0.50-1.62) | 0.87 (0.48-1.57) |
|  | 40-49 | 659 (34.8) | 66 (10.0) | 1.6 (1.3-2.1) | 0.92 (0.52-1.61) | 0.93 (0.53-1.63) |
|  | 50-59 | 394 (20.8) | 51 (12.9) | 2.2 (1.7-2.9) | 1.21 (0.68-2.15) | 1.20 (0.67-2.15) |
|  | ≥60 | 226 (12.0) | 24 (10.6) | 1.8 (1.2-2.6) | 0.92 (0.52-1.87) | 1.00 (0.52-1.92) |
|  | missing | 2 (0.1) |  |  |  |  |
| **Nationality** |  |  |  |  |  |  |
|  | Portuguese | 1824 (96.4) | 192 (10.5) | 1.8 (1.5-2.0) | ref. | ref. |
|  | Non-Portuguese | 65 (3.4) | 8 (12.3) | 2.1 (1.1-4.3) | 1.22 (0.60-2.47) | 1.38 (0.67-2.80) |
|  | missing | 2 (0.1) |  |  |  |  |
| **Comorbidities <sup>d</sup>** |  |  |  |  |  |  |
|  | No | 1141 (60.3) | 117 (10.3) | 1.7 (1.5-2.1) | ref. | ref. |
|  | Yes | 746 (39.5) | 84 (11.3) | 1.8 (1.5-2.3) | 1.06 (0.80-1.41) | 1.00 (0.74-1.34) |
|  | missing | 4 (0.2) |  |  |  |  |
| **Educational level** |  |  |  |  |  |  |
|  | Doctorate | 813 (43.0) | 83 (10.2) | 1.7 (1.4-2.1) | ref. | ref. |
|  | Bachelor's or master's degree | 797 (42.1) | 81 (10.2) | 1.7 (1.4-2.1) | 1.00 (0.73-1.36) | 1.01 (0.73-1.40) |
|  | Secondary or post-secondary | 187 (9.9) | 22 (11.8) | 1.9 (1.3-3.0) | 1.14 (0.71-1.83) | 1.10 (0.68-1.76) |
|  | Basic | 93 (4.9) | 15 (16.1) | 2.6 (1.6-4.4) | 1.55 (0.89-2.68) | 1.42 (0.81-2.48) |
|  | missing | 1 (0.1) |  |  |  |  |
| **High-skilled white-collar** |  |  |  |  |  |  |
|  | No | 305 (16.1) | 44 (14.4) | 2.5 (1.8-3.2) | ref. | ref. |
|  | Yes | 1579 (83.5) | 156 (9.9) | 1.6 (1.4-1.9) | 0.69 (0.49-0.96) | 0.73 (0.52-1.02) |
|  | missing | 7 (0.4) |  |  |  |  |
| **Remote working at the time of the second evaluation** |  |  |  |  |  |  |
|  | No | 763 (40.3) | 91 (11.9) | 2.0 (1.6-2.5) | ref. | ref. |
|  | Yes, part-time | 881 (46.6) | 83 (9.4) | 1.6 (1.3-1.9) | 0.77 (0.57-1.04) | 0.78 (0.57-1.05) |
|  | Yes, full-time | 224 (11.8) | 22 (9.8) | 1.6 (1.0-2.4) | 0.77 (0.48-1.24) | 0.82 (0.51-1.33) |
|  | Does not apply <sup>e</sup> | 21 (1.1) | 4 (19.0) | 3.5 (1.3-9.4) | 1.74 (0.64-4.73) | 1.65 (0.60-4.53) |
|  | missing | 2 (0.1) |  |  |  |  |
| **Area of residence (NUTS 3)** |  |  |  |  |  |  |

|  |  |  |  |  |  |
| --- | --- | --- | --- | --- | --- |
| Outside MAP <sup>f</sup> | 142 (7.5) | 9 (6.3) | 1.1 (0.6-2.0) | ref. | ref. |
| MAP | 1748 (92.4) | 193 (11.0) | 1.8 (1.6-2.1) | 1.74 (0.89-3.39) | 1.58 (0.80-3.11) |
| missing | 1 (0.1) |  |  |  |  |
| **Self-perceived probability of infection** | **(n=1822)** | **(n=135)** |  |  |  |
| Very low or low | 1192 (65.4) | 84 (7.0) | 1.2 (1.0-1.5) | ref. | ref. |
| Moderate | 527 (28.9) | 41 (7.8) | 1.3 (1.0-1.8) | 1.10 (0.76-1.60) | 1.14 (0.78-1.66) |
| High or very high | 103 (5.7) | 10 (9.7) | 1.6 (0.9-3.1) | 1.39 (0.72-2.68) | 1.45 (0.75-2.80) |

aIRR, adjusted Incidence Rate Ratio; CI, Confidence Interval; IR, Incidence Rate; IRR, Incidence Rate Ratio; MAP, Metropolitan area of Porto; NUTS 3, Nomenclature of Territorial Units for Statistics Level 3.
a Incident cases were defined as having a positive result in the second evaluation after having a negative in the first, among those who participated in both evaluations;
b Cases may not sum up 202 due to missing data;
c Adjusted for sex, age (continuous), nationality, and occupation classification; Education level was not adjusted for occupation;
d Comorbidities were defined as having a disease that requires regular medical care (e.g. treatments, appointments, etc);
e Does not apply, means being in parental, sabbatical, or sick leave;
f Outside MAP: participants' residence in Cávado, Aveiro Region, Tâmega and Sousa, Alto Minho, Alto Tâmega, Metropolitan Area of Lisbon, Douro, Coimbra Region and Viseu Dão Lafões;
\*Fisher exact test.

Table 2 shows the clinical and infection-related characteristics of workers that were seropositive at the first evaluation (n=69) and newly seropositive at the second (n=202). In brief, 82.6% and 46.0% never had a molecular test, respectively. From a clinical perspective, 29.0% and 9.4% were asymptomatic; 14.5% and 33.3% reported a known contact with a case and, of those, 60.0% and 69.2% were quarantined, respectively.

**Table 2.**
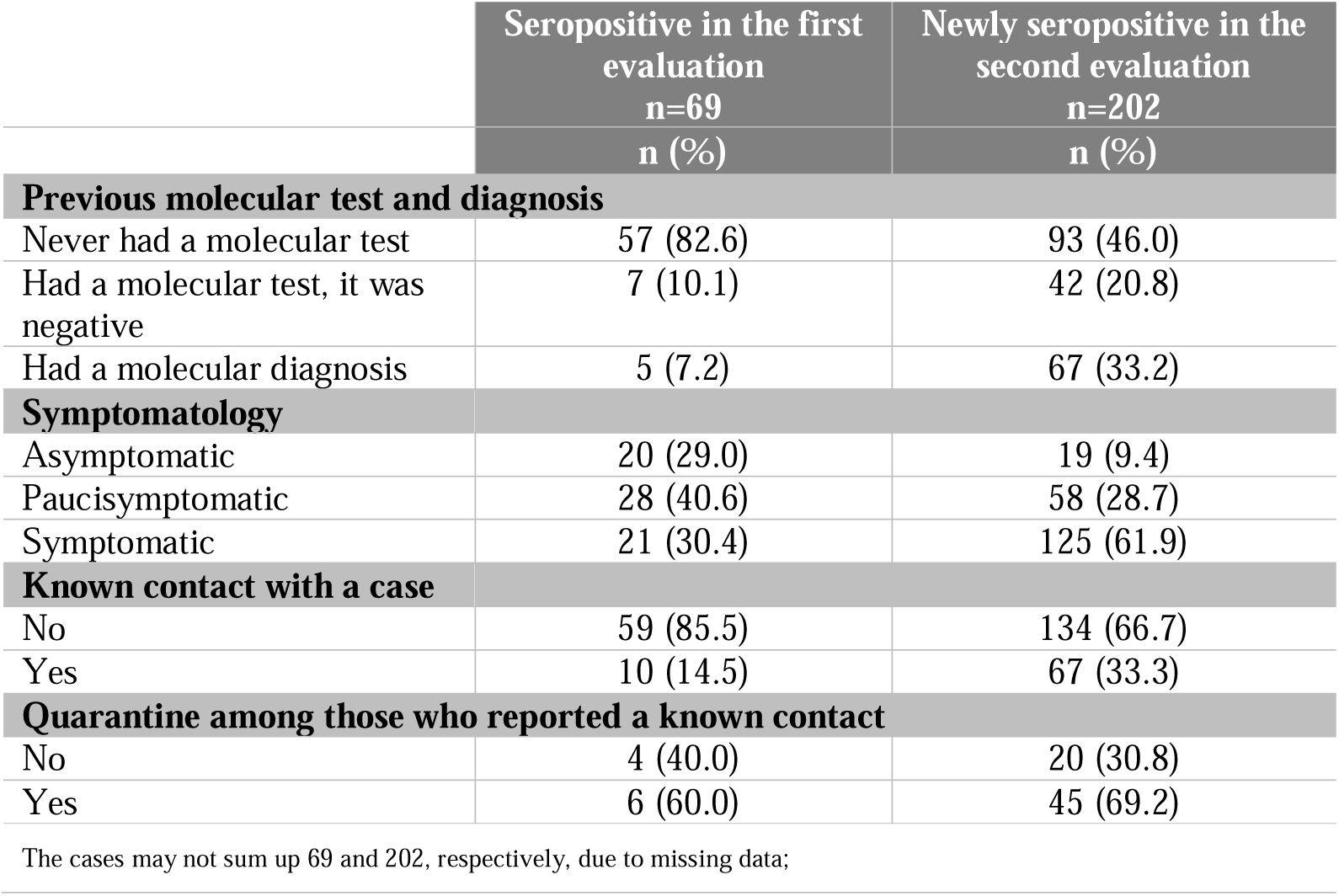
Clinical and infection-related characteristics of workers that were seropositive in the first evaluation (n=69) and newly seropositive at the second (n=202).

|  | Seropositive in the first<br>evaluation<br>n=69<br>n (%) | Newly seropositive in the<br>second evaluation<br>n=202<br>n (%) |
| --- | --- | --- |
| **Previous molecular test and diagnosis** |  |  |
| Never had a molecular test | 57 (82.6) | 93 (46.0) |
| Had a molecular test, it was<br>negative | 7 (10.1) | 42 (20.8) |
| Had a molecular diagnosis | 5 (7.2) | 67 (33.2) |
| **Symptomatology** |  |  |
| Asymptomatic | 20 (29.0) | 19 (9.4) |
| Paucisymptomatic | 28 (40.6) | 58 (28.7) |
| Symptomatic | 21 (30.4) | 125 (61.9) |
| **Known contact with a case** |  |  |
| No | 59 (85.5) | 134 (66.7) |
| Yes | 10 (14.5) | 67 (33.3) |
| **Quarantine among those who reported a known contact** |  |  |
| No | 4 (40.0) | 20 (30.8) |
| Yes | 6 (60.0) | 45 (69.2) |

The cases may not sum up 69 and 202, respectively, due to missing data;

## Discussion

In this cohort of university workers, the incidence rate was 1.8 infections per 100 person-months, and the 6 months’ cumulative incidence was 10.7%. The frequency of infection was higher based on seropositivity than molecular diagnosis, similarly to what has been described in most seroprevalence studies (3–9). However, the ratio serological/molecular diagnosis of SARS-CoV-2 infection markedly decreased with time, from 10:1 in the first evaluation to 3:1 in the second. This difference might be due to the increasing capacity of testing over the epidemic, as the proportion of individuals that were seropositive and never had a molecular test decreased from 82.6% at the first evaluation to 46.0% at the second evaluation. Similarly, the second round of the ISN COVID-19 also found a lower ratio between seropositivity and reported molecular diagnosis than during the first round (17). However, these differences might be overestimated due to the lower frequency of infection over the first months of the epidemic (18).

These results stress that we miss many infections making public health efforts less efficient. The differences in seroprevalence and cumulative incidence were thought to be mainly due to asymptomatic infections, however, in our study, most of those who were seropositive without a previous molecular diagnosis were classified as pauci- or symptomatic considering the reported symptoms, and this is in accordance with an earlier study (3). Moreover, most of them did not report a known contact with a confirmed case, then they were not likely missed by the contact tracing system. Similarly, in the Liverpool population-wide asymptomatic rapid antigen testing program, testing uptake was lower among populations with higher positivity (19) and another study showed that undetected infections were more frequent among those lower educated (20). Efforts must be done to identify and address the barriers to test-trace-isolate, particularly when COVID-19 like symptoms are present.

As previously shown less educated and less skilled workers were more likely to be seropositive and to get infected during the observation period (20–22), although our participants had an average high level of education and worked in a favoured context, those less educated and lower-skilled were still at higher risk of infection. However, we were not able to assess if less educated or less skilled workers were more likely infected due to overall social disadvantages, occupational exposure, household exposure, or because preventive messages were not conveyed considering the different literacy levels of the target population.

We found lower seroprevalence among those who live outside the metropolitan area, mostly in lower density populations, in accordance with findings of the nationwide seroepidemiological study conducted in Spain, with higher seroprevalence in more populated municipalities (6). We found a higher incidence among females, contrasting to the ISNCOVID-19 results and the population-based study in Geneva (4, 5), while no differences were found in Spain (6) and the Netherlands (8). In our sample, females had more frequently lower-skilled occupations and lower education. The Geneva study found a lower risk of being seropositive among those aged 65 years or more compared to those aged 20-49 years (5, 23). We found no statistically significant differences according to age, probably reflecting the homogeneity of the population studied, mostly higher educated and still working participants.

Although we have no data on workers affiliated at U.Porto whose salary comes from external financing sources we do have data on those hired by U.Porto and their distribution based on sociodemographic characteristics is similar to the observed in our sample. By the end of December 2020, there were 4798 workers, 2584 (54%) were females, 2238 (46.6%) had a Ph.D., 1894 (39.5%) a bachelor or master’s degree, 450 (9.4%) secondary education, and 216 (4.5%) basic education (24).

We found that 42.9% of the workers were IgM-only reactive at the two serosurveys, suggesting that they remained with detectable levels of IgM over the mean six months of follow-up, at least. Although the levels of IgM appear to decay 4-5 weeks after symptom onset, some patients remain positive at least 3 months after (25–27). However, it is not clear if they developed IgG or if they developed in undetectable amounts or either for a short period. We cannot fully explain why 38.9% of the workers who were IgG-only reactive in first were IgG and IgM reactive in the second evaluation, it may be either due to false negative or false positive results, once that the IgM usually peaks earlier (27).

This study has some limitations, as workers were self-selected, and the invitation was sent by e-mail. We sought symptoms, contacts, and episodes of quarantine since January 2020; they all were self-reported, but these questions are not expected to be prone to social desirability. However, as SARS-CoV-2 infection symptoms are unspecific they may be prone to recall bias. We completed the questionnaire before sharing the serological results with the participants, to avoid contamination by the rapid test result. This study is one of the few internationally focusing on higher education workers, a work environment where infection awareness is expected to be very high, and one of the few with a longitudinal approach.

To conclude, we found that university workers, in Porto, presented a high incidence rate of infection during a period of restrictive measures (2 infections per 100 person-month and a 6 months’ cumulative incidence of 10.7%), with the incidence being lower in males and “high-skilled white-collar” workers. The frequency of infection based on the serological tests was much higher than based on molecular diagnosis data, although it decreased over time reflecting wider access to testing. Nonetheless, these results stress that we still miss opportunities to test-trace-isolate persons with the infection, particularly those with symptoms but no known contacts, even in a work environment where infection awareness is expected to be very high.

## Data Availability

The dataset analysed during the current study is available on reasonable request to the study coordinator (PM).

## Author contributions

JPC participated in the study design, wrote the draft of the manuscript, and performed data analysis. PM and HB designed the study and provided guidance for data analysis and interpretation of results. PM, PNSR, and HB critically reviewed the draft. All authors revised and approved the final version of the manuscript for submission.

## Acknowledgements

We wish to acknowledge the team of researchers in the field. IT support from Paulo Oliveira. The health professionals from the Occupational Service, Infectious Diseases Service, and the Clinical Pathology Service from University Hospital Center São João. This study was funded by the University of Porto and supported by national funds of *Fundação para a Ciência e Tecnologia* (FCT), under the scope of the project UIDB/04750/2020 - Research Unit of Epidemiology–Institute of Public Health of the University of Porto (EPIUnit). JPC is the recipient of a Ph.D. grant (DFA/BD/8562/2020) co-funded by the national funds of FCT and the *Fundo Social Europeu* (FSE). Authors declare no conflicts of interest.

## Notes

### Competing Interest Statement

The authors have declared no competing interest.

### Funding Statement

This study was funded by the University of Porto and supported by national funds of Fundacao para a Ciencia e Tecnologia (FCT), under the scope of the project UIDB/04750/2020 - Research Unit of Epidemiology-Institute of Public Health of the University of Porto (EPIUnit). JPC is the recipient of a Ph.D. grant (DFA/BD/8562/2020) co-funded by the national funds of FCT and the Fundo Social Europeu (FSE). Authors declare no conflicts of interest.

### Author Declarations

The study protocol was approved by the ethics committee of the Institute of Public Health of the University of Porto (ID 20154).

