## Supplementary material for "Incidence of SARS-CoV-2 infection in a cohort of workers from the University of Porto": Table S1.

**Table S1.** Comparison of serological results among those who participated in both evaluations (n=1960).

|  |  | **Second Evaluation** | | | |  |
| --- | --- | --- | --- | --- | --- | --- |
|  |  | Seronegative | Only IgM | Only IgG | IgM and IgG | Total |
| **First Evaluation** | Seronegative | 1689 | 68 | 16 | 118 | 1891 |
|  | Only IgM | 18 | 21 | 0 | 10 | 49 |
|  | Only IgG | 2 | 0 | 9 | 7 | 18 |
|  | IgM and IgG | 0 | 0 | 1 | 1 | 2 |
|  | Total | 1709 | 89 | 26 | 136 | 1960 |
| Ig, Immunoglobulin. | | | | | | |
